# Walking the line between assessment, improvement, and learning: an exploratory study on opportunities and risks of peer discussion of audit and feedback

**DOI:** 10.1101/2022.07.19.22277796

**Authors:** Dorien van der Winden, Nynke van Dijk, Mechteld Visser, Jettie Bont

## Abstract

**Introduction:** There is a broad call for change in existing quality systems within health care. One of the anticipated reforms, is transferring ownership back to care deliverers. A promising way to establish this in general practice care, is to combine audit and feedback with peer group discussion. However, it is unknown what different stakeholder groups think of giving prominence to this type of quality improvement. In this study we explore ideas and opinions of different stakeholder groups in general practice on the opportunities and risks that could arise.

**Methods:** We conducted an exploratory qualitative study, combining interviews with focus discussion groups. Included stakeholder groups were general practitioners, patients, professional organizations and insurance companies. Within a constructivist paradigm, two researchers coded the data in three rounds, using thematic analysis. After continuously comparing and discussing codes with a third researcher, a final code tree emerged, presenting us with the main themes.

**Results:** In eight interviews and two focus discussion groups, 22 participants reflected upon opportunities and risks. We identified three main opportunities: deeper levels of reflection upon data, adding context to numbers and more ownership, and three main risks: handling of unwilling colleagues, lacking a safe group and the necessity of patient involvement. An additional theme concerned disagreement on the amount of transparency that should be offered: insurance companies and patients advocated for complete transparency on data and improvement of outcomes, while GPs and professional organizations urged to restrict transparency to giving insight into the process.

**Conclusion:** Peer discussion of audit and feedback could be part of a change movement, towards a quality system based on learning and trust, that is powered from within the profession. Creating a safe learning environment is key herein. Caution is needed when complete transparency is asked, since it can jeopardize practitioners’ reflection and learning in safety.

**Strengths and limitations of this study:**

- Exploratory study of a gap in the knowledge towards implementation of a promising intervention.
- Participants included the four main stakeholder groups that are involved in implementation, in order to study agreement and disagreement between different stakeholders.
- Additional stakeholders, such as governmental agencies, were not included. They may have offered other perspectives.
- This exploratory qualitative study offers insight into opportunities and risks. In order to get a broad overview of how these findings are supported by GPs in general, additional quantitative research is necessary.

## INTRODUCTION

In recent years, health care professionals have called for the reform of existing quality improvement structures, aiming to change the focus from systems based on accountability to systems based on trust and development. One way to reform current structures is by adding peer discussion to individual audit and feedback reports. In this paper we explore the ideas and opinions of stakeholders on this development.

Current quality systems were established in the 1980s by governments and other supervisory bodies in high-income countries, partly in reaction to increasing demands for transparency and accountability in health care.^1^ Over the years, the emphasis of these systems shifted to auditing of performance indicators.^2-4^ In the past decade, researchers and policy makers, as well as health care providers, have acknowledged that these structures have many disadvantages. These disadvantages include a high administrative burden and a possible decrease in motivation among professionals working within these systems.^5-7^ Although designed to assess and ensure high levels of care, systems often focus on inconsequential indicators and do not necessarily improve the quality of actual patient care.^8-10^

Doctors and other health care professionals want to reclaim ownership of quality measures, and stress that the focus must shift from assessment to significant improvement of patient care.^11 12^ Quality improvement researchers and policy makers support this move. For example, in a 2016 viewpoint in JAMA, Donald Berwick proposed that a new “era” for health care should now arise. This should be an era in which we let go of excessive measurement and transform into a “learning system.”^1^ In 2018 Jeffrey Braithwaite, laid out in BMJ that in order to sustain actual improvement in health care, a different mindset towards quality policy is necessary, appreciating a more nuanced form of quality improvement.^8^ Professional organizations for general practice in the Netherlands are currently advocating the development of a quality improvement system that focusses on collaborative learning and improvement. In their joint vision document on quality policy in general practice, published in 2019, they recommend increased use of peer-to-peer coaching and assessment, among other measures.^13^

A promising way to give peer coaching and assessment a vital role in quality improvement could be small-group peer discussion of audit and feedback (AF). This combines two notable forms of quality improvement measures in general practice: audit and feedback and small-group peer discussion. AF interventions are widely used in quality improvement. In these interventions, clinical practice is measured and summarized using indicators. The results are then communicated back to the health professionals, with the purpose of establishing reflection upon their practice.^14^ Research has shown that interventions based on AF have a positive, though mild, measurable effect on professional practice.^15 16^ Small-group peer meetings, in the form of quality circles, have become a major part of continuing professional development (CPD) and quality improvement in general practice.^17^

Current research on AF focusses on how to effectuate the best results with an AF intervention.^14 18-20^ As AF interventions are intended to change the behavior of the professionals concerned, the Behavior Change Wheel is increasingly used to offer insight into influencing factors. In this framework, Michie et al. explain that opportunity, motivation, and capability have a mutual influencing role when trying to change behavior.^21^ In small-group peer discussion of AF, a group of professionals review their individual data and develop an action plan to improve their practice. Previous research has shown that this way of reviewing AF reports with peers seems to heighten motivation to change and leads to change planning.^22 23^ Incorporating peer discussion in an AF intervention therefore seems to influence opportunity, motivation, and capability.

Although small-group peer discussion of AF reports seems promising, it is largely unknown what the stakeholders concerned in general practice think about giving this method a more prominent role in quality improvement. Insight into the opinions and ideas of stakeholders is indispensable in order to facilitate implementation and ensure uptake. In this study, we therefore spoke with stakeholders in general practice to explore the opportunities and risks of small-group peer discussion of AF. We asked them their views on peer discussion of AF as part of a new era of quality improvement and explored the role of transparency and accountability in such a system.

## METHODS

### Study design

For this qualitative study, a constructivist paradigm was adopted to explore views and ideas of different stakeholders who function in different professional contexts in general practice. Thematic analysis was used to identify patterns in these differing viewpoints.^24^

### Setting

This study was conducted in the Netherlands, in a general practice context. All inhabitants of the Netherlands are registered with a specific general practitioner (GP) practice, where they go for diagnosis and treatment of all initial symptoms, and/or referral if necessary. General practitioners (GPs) therefore have a strong gatekeeper function within the Dutch health care system.^25^ GPs have to renew their license every five years. This requires two hundred hours of CPD activities. At least ten of these hours must be dedicated to peer-to-peer activities, e.g. peer-to-peer coaching, feedback, or discussion of AF reports.^26^ Both professional organizations and insurance companies play a role in the quality system within general practice. Professional organizations advocate for GPs at national and regional levels with regard to quality policies. They also manage guideline development and many professional organizations provide CPD activities. Funding of GP surgeries is managed through insurance companies, which provide AF reports to GPs on an annual basis.^27^

### Participants

Participants were relevant stakeholders in the Dutch general practice setting: GPs, patients, representatives of professional organizations for GPs, and representatives of insurance companies. Purposive sampling was conducted in order to include stakeholders with different views. GPs and patients were recruited through the academic network of the general practice department of our university. We selected patients through the patient board of a large umbrella organization of GP practices, to make sure our participating patients had some understanding of the GP policy setting, in order to form an opinion on quality measures and continuing professional development of GPs.

### Data collection and analysis

Data collection and analysis took place using an iterative approach, from December 2019 until June 2020. We chose different data collection approaches for different stakeholders in order to achieve optimal conditions: homogenous focus discussions for GPs and patients, and individual interviews for the representatives of professional organizations and insurance companies. The homogenous focus group discussions with GPs and patients enabled discussion between participants, leading to clarification of individual viewpoints and revelation of mechanisms behind their ideas.^28^ The semi-structured in-depth interview design used for the representatives of professional organizations and insurance companies allowed them to provide more in-depth information on their thoughts and ideas, while preventing the appearance of a political meeting.^29^ Interview duration was between 45 and 60 minutes. Focus group discussions lasted 55 minutes for the GPs and 75 minutes for the patient group. Interviews were held at a locations that were convenient for the participants. Focus group discussions were held at central locations.

In order to obtain insights on the same themes, the same topic list was used to conduct both the focus group discussions and the interviews. We asked the participants for their opinions on four main themes: possible purposes of AF peer discussion meetings, how to incorporate these meetings in a quality system, the role of accountability and transparency, and how best to implement a quality system containing A&F peer discussion.

During the interviews and focus discussions, we used an infographic on how a simple system based on peer discussion of AF could be designed (Figure 1). This infographic was designed by the researchers based on preliminary conversations with different stakeholders. We used the infographic as the starting point of the conversations in order to clarify a complex system and to check whether ideas on the basic design of such a system aligned. We verified our findings through member checking of the results, by sending a summary to our participants. We aimed to achieve data sufficiency by including all different viewpoints.

**Figure 1.**
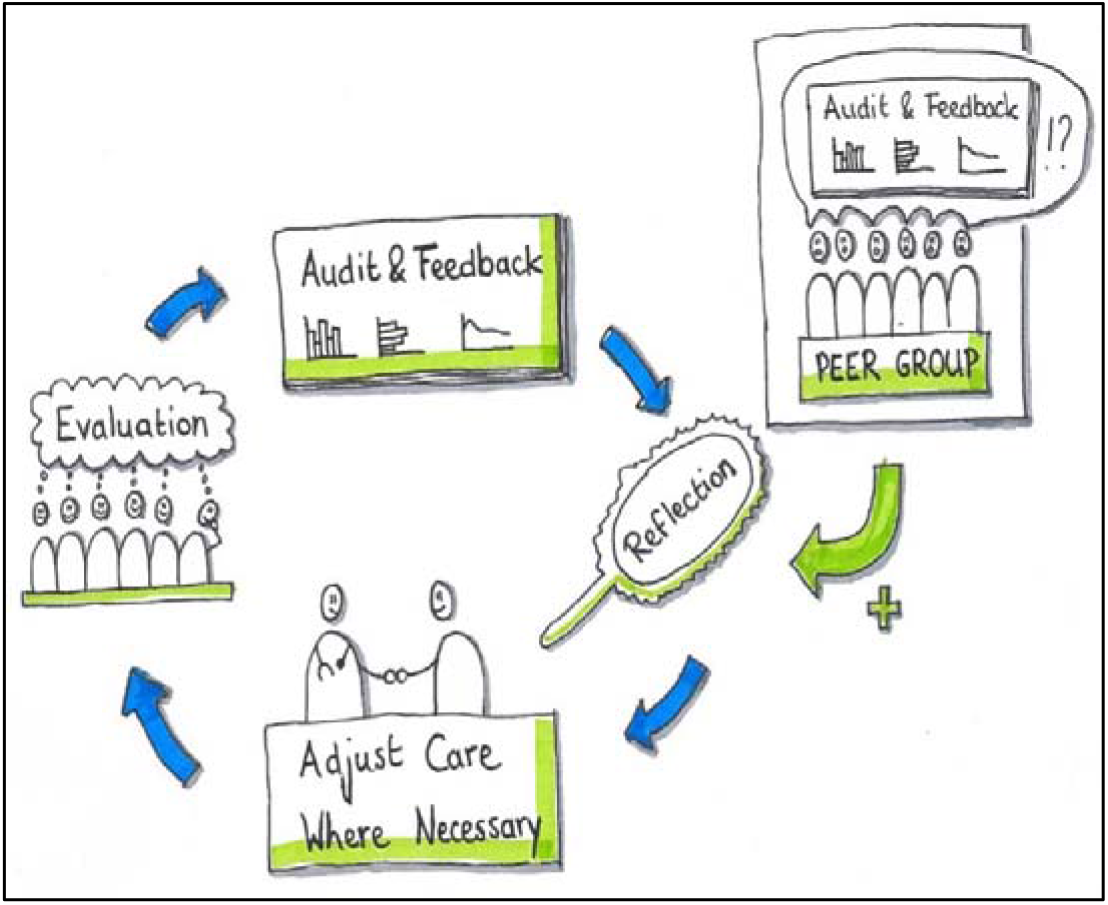
Infographic on group discussion of A&F

Interviews and focus group discussions were audiotaped, transcribed verbatim and anonymized. Transcripts were analyzed with a thematic analysis approach, using MAXQDA.^24 30^ The first three transcripts were analyzed by two researchers (DW and JB) using open coding. Next, the two code trees were compared and discussed in detail, resulting in one preliminary code tree. DW coded the remaining seven transcripts. Ambiguous fragments were discussed with JB until agreement on coding was reached. This resulted in the final code tree. DW, JB, and ND read through all the transcripts individually once more and discussed the code tree. From this discussion, the final themes were established by consensus of the full research team. We reported according to the SRQR-checklist for qualitative research.^31^

### Patient and public involvement

Patients were included as one of the stakeholder groups in general practice (see participants). The results of this study will be shared with all participants, including the patients.

### Reflexivity and ethics

The researchers are affiliated with a large general practice research and training institute. The research team included a GP trainee (DW), a GP and head of general practice department (JB), a cognitive psychologist (MV), and an MD/medical educator (ND). They all work as medical education researchers, some with a long history of research in general practice education (MV, ND) and are therefore familiar with the setting. The research project was prompted by a request made by a group of local GPs asking for a scientific framework for an alternative quality cycle in general practice. This request informed a larger research project of which this is the first exploratory study.

Throughout this project we have aimed to conduct reflective research, giving all viewpoints equal consideration. To prevent biased interpretation of data, we have kept a reflexive stance throughout the research process by gathering frequently to discuss our positionality and the implications thereof.

We received a waiver from the Medical Ethics Review Committee of the Amsterdam University Medical Centers, confirming that the Medical Research Involving Human Subjects Act does not apply to our study. Participation in this study was voluntary. We asked for and received informed consent from all our participants. All data obtained within this study was processed and stored in accordance with the General Data Protection Regulation and the Amsterdam UMC Clinical Research Unit procedures.

## RESULTS

We conducted eight interviews and two focus discussions with a total of 22 participants.(table 1) We asked our participants for their opinions on AF peer discussion and its positioning within the quality system of general practice. The exploratory nature of this study resulted in rich data. We therefore chose to focus on the main themes that arose around the foundations of peer discussion of AF, in alignment with our research question. The three main themes that emerged were “what are the opportunities of peer discussion of A&F?”, “What are the risks?”, and “Disagreement on the amount of transparency.” These themes comprised different subthemes.(box 1 and 2) The full final code tree can be found in the supplemental materials.

**Table 1.** Characteristics of participants

| Stakeholder group | No of participants | Description |
| --- | --- | --- |
| GPs | 11 | Working in different types of practices (7 in solo, 4 in dual/group practices)<br>Working in different areas (2 in rural, 3 in small-town, 6 in urban areas) |
| Professional organizations | 4 | Board members of different POs |
| Insurance companies | 3 | Representatives with different functions within the organization<br>Differing from board member to medical adviser |
| Patients | 4 | Representatives of a patient board of a large GP organisation |
| Total | 22 |  |

### What are the opportunities of peer discussion of AF

Participants identified several opportunities that could be offered by peer discussion of AF. They talked about what the group process can offer in addition to looking at AF reports individually. The three main opportunities they mentioned are: reaching deeper levels of reflection, adding context to numbers, and taking more ownership of quality improvement.

#### Box 1

**Opportunities**

**<u>Opportunities of peer discussion of AF</u>**

Reaching deeper levels of reflection

Adding context to numbers

More ownership of quality measures lies with the GPs

#### Reaching deeper levels of reflection on daily practice

All participants agreed that discussing AF reports with peers deepens reflection on daily practice, compared to reflecting on these reports on one’s own. The data serves as the first mirror to which your practice is held up. The group acts as a second mirror, as one of the participants pointed out. According to the stakeholder groups, the group deepens personal reflection by asking questions participants would not ask themselves. It also helps to uncover blind spots, shows you solutions you would not have found on your own, motivates you to actually change your behavior, and encourages you to stick to your improvement plan. Participating patients also explicitly called for GPs to work together when it comes to quality improvement.(see box 2 for quotes)

##### Box 2

**Reaching deeper levels of reflection**

*“You have a second mirror. There’s a second mirror here ¡polnis to the group in the diagram]. This is the first mirror with information [points ta theA&F report]. The second mirror is what your rnllengue says about you in that respect, ar perhaps what they soy nhnut your reflection That has real added value, of course.” (PC2)*

*“Then you talk to your colleagues, and ask them ‘hey, how would you answer that question,’ And then you say ‘hey, there’s another way after all ⃛ ‘They’re often very obvious things that make you think ‘oh. gosh, It can be done that way too. It’s a blind spot*.*’” (GP1)*

*“You don’t necessarily hove to, but i think that doing It in a group takes the mirroring further. If i just look at a few figures and see ‘t’m doing those things well, bin I’m not doing those quite as well, i’ll need to look at those again,’ there’s a good chance it wilt stop there. There {points to tne group) you’re encouraged to think about it more and answer questions like ‘what are you going to do with it, how, why and when?’” (PO3)*

*Pt 1* ; *“That’s exactly whet you don’t want anymore, for the GP to do it all an his Own, but for him to… I think a group like this is essential. … Yes, it should he required.” (Focus discussion 2)*

#### Adding context to numbers

GPs and representatives of professional organizations mentioned that peer discussion helps to solve the perceived oversimplicity of A&F reports. They expressed being frustrated with how outcome reports are often used in quality improvement: being assessed by numbers that simplify the complex reality of their patients and of the patient centered care they provide feels unjust. However, A&F reports become more meaningful when used differently: as a basis for deeper reflection on practice, thereby prompting a conversation on improvement.

Participants among all stakeholder groups agreed that peer group discussion therefore adds meaning and explanations to the indicators, doing justice to the complexity of general practice care.(box 3)

##### Box 2

**Adding context to numbers**

*“If you’re in a group, of course you can always talk to each other about it. That’s another added value, because then you can also look at what that average says’ or bow are we all doing?’” (PO1)*

*“The sum of all that mirror information is really useful. Not in the absolute sense of basing a judgment on it, butin the sense that it can encourage you to reflect on your own performance*.*”* ***(P04)***

*“If I may say so, an indicator is nothing more than what the word says: an indicator. As far as I’m concerned, it’s simply an invitation for a discussion, in terms of ‘reflection, self-reflection*.*’” (IC3)*

#### Taking more ownership of quality improvement

Discussing audit and feedback in a group of fellow GPs was seen as a way to take back ownership of quality measures in general practice care. The GPs expressed hope that introducing a quality system whose cornerstone is peer discussion of A&F will lead to a next-level quality system in which professionals are constantly learning with and from each other. Representatives of the professional organizations were of the same mind. The GPs and representatives of professional organizations expressed a desire for a system powered from within the profession, resulting in less externally imposed standards and more meaningful quality improvement. Representatives of insurance companies supported this transfer of ownership back to the professionals. GPs explained that ownership over quality improvement is also increased through the group process in another way: when a shared difficulty is identified, a group of professionals has more power to change the context they are working in, as compared to the individual GP.(box 4)

##### Box 4

**Taking more ownership**

*“Well, you know? It stimulates the strengthening of the intrinsic motivation of GPs to work on quality, within yourself and within the group. It prevents you from having to constantly account fcr your actions externally. That’s the whole train of thought behind it. So that’s why I think it’s a good idea.” (P02)*

*“Well, of course I’m really happy that there are parties wno say ‘i wont It (the power over their own quality policy] back.’ I think it’s perfectly normal to evaluate your actions ps a doctor. That’s part of vour medical professionalism.” (¡ C1)*

*“If you all conclude that ‘there’s something wrong with our context,’ that you’re then strong enough to take it to the next level together, that ‘something really has to change,’ instead of always fighting it out nt the practical level and then often not tackling it in depth.”*(GP1)

### What are the risks

Having considered the opportunities, participants also mention several risks that could occur when making peer discussion of AF the cornerstone of your quality improvement system. Because this system relies heavily on the willingness of GPs to improve practice, the question of how to handle unwilling colleagues arose. GPs also mentioned the necessity of having a safe group of peers in which to participate. Patients and GPs raised the subject of patient involvement and how important it is to incorporate this into the system.

#### Box 5

**Risks**

**<u>Risks of peer discussion of AF</u>**

How to handle unwilling colleagues?

Lacking a safe group

Patients should he involved

#### How to handle unwilling colleagues

Participating GPs mentioned that, although they believe the majority of their colleagues will be eager to participate, there will always be peers who are not motivated to participate and/or change their practice, even with the best reflection methods. Participating GPs seem to accept this as a given, and conclude that this cannot be overcome by any quality system. Participants stated that this should be the responsibility of the colleagues within the group and of the regional organizations. However, one of the GPs pointed out that those doctors who are not keen to participate and change their practice in this way could still be excellent doctors for their patients. Another representative of a professional organization explained that there are already measures in place to ensure patient safety, such as regulations concerning license renewal and Healthcare Inspectorate.(box 6)

##### Box 6

**Unwilking colleague**

*“Yes, personally t believe that colleagues play the greatest role in this. That’s the best thing, because they kne w. We actually know the real people who make a mess of things. I know them. Everyone knows them. You get to know them after a while. Or the people with problems, their own problem, who therefore perform inadequately. Of course, the trick is for the sector itself to have a certain self-cleansing capacity as well…. i think that that [holding each other accouniabie for inadequate performance] happens occasionally. I can’t really judge how often, i think it certainly does happer., but I also think It often doesn’t happen. That’s something we need to get better at, I think” (PO4)*

*“if you have a colleague who really performs inadequately and ng real change occurs, then of course we ourselves also have a responsibility todo something about it, also with respect to the ‘nspectorate perhaps. But then again, not everyone is equally good ar does everything equally well. You’re not simply going to report colleagues who aren’t doing so well in a group like that. Such a group isn’t suitable for that and really isn’t intended for that either.” (PO1)*

#### Lacking a safe group

All stakeholders indicated that “feeling safe” is an important prerequisite for reflection to actually take place. Participants agreed that many GPs in the Netherlands have a safe peer group in which they participate. However, participants mentioned that there are also groups of GPs lacking mutual trust, even while practicing their obligatory peer-to-peer CPD activities together. This could be caused by bringing groups together due to geographical location. Competition for patients can be an issue in these groups, resulting in an environment in which GPs do not feel safe to freely reflect upon their work. Participants expressed that this could impair learning.(box 7)

##### Box 7

**Lacking a safe group**

*“An assessment group is fine if you have a group in which you trust each other, where there is an obligation of confidentiality, and where you can therefore assess yourself and be assessed. But that’s not the same in every GP group. That’s not always the same as the CPD group*.*” (GP11)*

#### Patients should be involved

Both GPs and patients mentioned that it is vital to involve patients in the AF peer discussion quality improvement cycle. Patient satisfaction often plays no part in current AF reports. GPs preferred to see patient satisfaction as a major indicator, since it says a lot about a practice: participants saw it as the most important “outcome” of their work as a GP. Participants in the patient focus discussion also favored patient involvement and suggested that patients could have a role in determining the subject of the AF, so that they could put matters that affect them the most on the agenda.(box 8)

##### Box 8

**Patients should be involved**

*Pt4: “I miss the patients in this whole circle*.*”*

*Pt 2. “Yes, I really miss them too”*

*Pt 3. “Then I would rather want to consult the patients of the peer group end ask ‘how do You feet tjbout this?’ I think GPs can learn more from that, also because I know from research that the stories behind the numbers say much more about the numbers than just the numbers,” (focus discussion 2)*

*“*… *ultimately, it’s about the information the patient gives back to us and we should be collecting nformation front the patient to see how our qua’ity of care is*.*(GP9)*

*“I think that if GPs decide for themselves, you’ll end up with their favorite topics and the loudest one will decide what happens. Perhaps you could also work with some sort of patient focus group, and ask ‘sc, we have ten topics now, what do you think is important?’ That it doesn’t just come from the doctors, because they might hove other interests than what is ultimately important for the patient group*.*”(Pt2)*

### Disagreement on the amount of transparency

Although all participants agreed that ownership of quality improvement should initially lie with the professionals themselves, issues were raised regarding the need for transparency of this process and/or its outcomes, in order to ensure accountability. Some of our participants argued in favor of process evaluation. Others stressed that this does not suffice and that some insight into outcome measurements is necessary. Patients showed ambiguity in their preferences on the amount of transparency that should be offered.

#### The argument for process evaluation

Professional organizations and GPs recognize that there is a need for some form of transparency on the quality of care that GPs provide. GPs and representatives of professional organizations share the view that this transparency should be offered in the form of process evaluation. They believe it should be sufficient to show the outside world through their mandatory annual report that they participate in AF peer group discussions in general: society should grant them “justified trust” when it comes to the results of the quality cycle. Opinions differ on whether to add what topics are being worked on and offering insight into the process. There is consent among the GPs and among most representatives of professional organizations that the amount of disclosure should be decided by the individual GP: it is seen as positive to share what is being worked on, but how much of it to share should be decided by the GP. An argument that our participants made in favor of this concerns the tension that can exist between an imposed level of transparency and the depth of reflection: if you need to be completely transparent, you do not feel truly free to make mistakes and reflect upon them.(box 9)

##### Box 9

**Tha argument for process evaluation**

*“So not just sitting in your ivory tower, but working on it with other people; I think that that should already show the outside world that we’re working hard on quality and that this can help the outside world get a better impression of the quality of GPs* ***themselves again*.*” (P01)***

*GP1: “Justified trust. Yes. That’s what it should be about, but that we as participants in such a group are really responsible together for ensuring that everyone actually puts their best foot* ***forward and comes with the intention of taking something away with them*.*”***

*GP7: “If that’s how you’re going to do it, based on justified trust and things like that, you can also leave a lot of control behind. Or it could be ‘never mind, you can read all about it in our annual* ***report’” (Focus discussion 1)***

*“The intrinsic part I think is the intrinsic quality perspective of the professional and the transparency discussion. What you sometimes see is that those two things get mixed up. That’s something we should try to avoid, because from that* ***intrinsic quality perspective, as a*** *professional you should actually be completely free to say ‘oops’ occasionally, to think ‘shoot, I* ***could have done that a bit better*’ *once in a while*.*” (IC3)***

#### Insight into outcome measurements is necessary

Representatives of the insurance companies and a representative of one of the professional organizations voiced that simple trust in the doctor to disclose the genuine weaknesses of his or her practice does not suffice in this day and age. Several representatives of the insurance companies opposed the fact that process evaluation should be enough: one would always represent oneself as functioning perfectly, or only offer insight into the things that improved, but not into the goals that were not achieved. For the latter, plain numbers are believed to be necessary.(box 10)

##### Box 10

**Insight into outcome measurements**

*“The problem is, everyore’s going to write something down, making it seem that everyone is doing great. it never means that much te me. I don’t know anyone who honestly writes down ‘we did this terribly, it’s still terrible, we failed. ‘ There’s too little trust for me. Then I would also like to see the hard data*.*” (IC1)*

#### Ambiguity in patient preference

The participants in the patient focus discussion were ambiguous regarding the necessity for transparency towards patients. They felt that, when it comes to medical technical skills, proper quality of care should be evident: as a patient you should be able to trust upon this without needing insight into numbers and outcomes. On the other hand, the participants acknowledged that some GPs may be better at certain things than others: you should be able to review whether your GP fits the bill on the issues you find essential. That may sway your decision to switch to a different GP, if geographically possible. If AF reports for their GPs were available, some of our participants would make use of them, provided that the numbers and measures were easy enough to understand. Even so, they stressed that their own experience of the quality of their GP remains the most important factor in determining whether they are satisfied, regardless of the objective measures into which they might have insight.(box 11)

##### Box 11

**Amtiguity in patient preference**

*“And it could be important for patients, because a patient would never choose a doctor who scored a 5 or 6 of course, but I think it’s especially important — in relation to education and perhaps follow-up courses — that you know how you score as a doctor, that you have areas for improvement based on that score. Ultimately that’s what it’s about.” (Pt2)*

*“So if I feel that my GP is competent, i’ll stay with my GP. if I have a GP, as I have had in the past, who is not competent, or a stand-in, which almost becomes a matter of life and death, I’ll never go there again and I’ll never want to see that stand-in again either. So that’s my own personci barometer, which is basically what you’re saying.” (Pt4)*

## DISCUSSION

In this study, we spoke to stakeholders in general practice to explore the perceived opportunities and risks of incorporating small-group peer discussion of AF reports in the changing quality improvement system in general practice. We identified several opportunities that peer discussion of AF could offer, encountered some risks, and discovered that there is disagreement on the amount of transparency that should be offered.

The opportunities that our participants described are: deepening of the level of reflection, addition of context to the numbers, and transfer of ownership of quality improvement to the GPs. The risks that we identified were: some GPs might be unwilling to participate or change, proper reflection occurs only in a safe group of peers, and it is important to add patient feedback to an AF cycle. When it comes to the role of transparency and accountability, there is disagreement between different stakeholder groups: GPs argue in favor of insight in the form of process evaluation, insurance companies state that they require at least some transparency on an outcome level, while patients show ambiguity in their preference.

## Opportunities

From our results it seems that AF reports and small-group peer discussion complement and reinforce each other when used together. By deepening reflection, peer discussion of AF boosts learning from AF and working towards change. By adding context to outcome measurements and transferring ownership of quality improvement to health care professionals, peer discussion of AF offers a partial solution to changing the quality system for the better, as called for by health professionals and researchers.

### Social learning, changing behavior, and feedback and assessment “for learning”

Many of our findings concerning the perceived opportunities of AF peer discussion tie in with existing medical educational literature: the idea that learning with a group of peers deepens reflection, heightens motivation, and increases ownership can be incorporated into medical and general educational theory. With his Social Learning Theory in the 1960s, Bandura introduced the notion that our learning occurs through the observation of others and is thereby a social process.^32^ Lave and Wenger later introduced the importance of communities of practice when it comes to professional development and learning: a group of peers sharing practices and experiences leads to enhancement of knowledge.^33^ Peer discussion of AF stands on the principles of these theories: a group of GP peers forms a community of practice. Within such a community, you not only learn from your own experiences, but also from seeing and hearing others. Sharing these experiences deepens reflection and thereby learning, as our participants confirmed to be the case with peer discussion of AF.

Our participants described that peer discussion deepens reflection on AF. It can therefore give meaning and momentum to the AF report, which are necessary for it to lead to improvements. When looking at the Behavior Change Wheel, a framework that provides insight into why and how people change behavior, peer discussion on AF could add to motivation to change and increase capability (two cornerstones of the Behavior Change Wheel), for example by sharing best practices and increasing ownership and capability to tackle shared problems.^21 34^

Additionally, peer discussion of AF fits within the four-step process proposed by Sargeant et al. in 2013 for using feedback and assessment “for learning”, rather than feedback and assessment “of learning”. ^35^ In their article on how feedback and assessment can encourage professional development, they explain that, as a first step, external data is necessary in order to improve practice, since self-assessment is not sufficiently reliable. With peer review of AF, this comes in the form of the AF report. The second step is engaging with the feedback. Sargeant et al. propose discussion of feedback in order to stimulate this. Discussing feedback leads to alignment of external feedback with the self-image and increases self-efficacy: it enables doctors to form an action plan. This aligns with the deeper levels of reflection described that peer discussion of AF provides, as described by our participants.

Peer discussion of AF therefore follows the long-standing rules of social learning theory. It appears to tie in with providing feedback “for learning,” engaging with it, and working towards behavior change.

#### Moving towards a learning quality system

When considering the other opportunities our participants expressed, it seems that giving peer discussion of AF a prominent role in the quality system of general practice could offer a partial solution to the problems that current quality systems showcase. Our participants believe that it can help solve the problem of losing context when looking solely at outcome measurements and that it transfers ownership of quality policies to the GPs. By being not only a quality improvement intervention, but also a CPD activity, we believe it will put the focus on learning and improvement instead of assessment. It would tackle some of the changes that Braithwaite proposed in 2018, which are necessary to change health care improvement.^8^ For example, it is powered from within the health profession (instead of top-down), it centralizes natural networks of clinicians, it pays attention to context, it does not solely focus on what went wrong, but also on what clinicians do right (by sharing best practices) and it is built on (and stimulates) collaboration. Furthermore, group discussion of AF fits into the new “era for Healthcare and Medicine”, that Berwick advocated for in his article in 2016: it will bring us closer to a “learning system”.^1^ Group discussion of AF takes into account the complexity of the environments in which GPs function, by giving them and their peers ownership of which subjects to act on, and how to act on them. It gives GPs the opportunity to focus on the measurements that matter.

### Risks

Berwick ends his article with the notion that stepping into this new era is not as easy as it seems.^1^ The risks our participants identified affirm this notion. The unresolved issue of what to do with GPs who are not willing to participate, meaningfully raises the question of whether this type of intervention is fit for accountability purposes. Berwick states that we should include and empower not only clinicians, but especially patients. As the patients and GPs in our study explained: the patient voice still needs attention within the peer discussion of AF cycle. The described necessity of a safe learning environment (relying both on having a trusted peer group and control over who has insight into the process and the outcomes) fits with educational theories on social learning. Even so, this safe learning environment clashes with the complete transparency that Berwick proposes to be necessary: participating GPs and professional organizations clearly argue for process evaluation, rather than outcome measurement.^1^

#### Transparency versus a safe learning environment

Importantly, disagreement on the level of transparency between our stakeholder groups presents us with a pivotal dilemma. While transparency is an important prerequisite when it comes to quality assessment, high demands for transparency may put the learning of health professionals, and thus quality improvement, at risk.

As the knowledge of quality improvement evolves, boundaries between quality assessment, improvement, and also CPD get blurred: we need to hold our health professionals accountable for what they do. We want them to improve their practice and we want them to learn and keep learning. All the while, we require them to show us not only how they are doing this, but also, to prove to us that it is working: we ask them to provide insight into both their actions and results. While this development has helped transform health workers into accountable professionals,^1^ going overboard with it puts learning, and thus sustainable improvement of practice, at risk.

This risk becomes clear when we revisit the previously discussed learning theories: a prerequisite for an individual to learn, and thus to improve practice, is a safe learning environment.^32 33^ If we look at peer discussion of AF, the necessity of a safe peer group is, as expressed by our participants, a given. Yet, the safety of the learning environment is not influenced only by the direct peer group, but also by others, such as insurance companies, Healthcare Inspectorate and patients, possibly looking over the doctors’ shoulders, at outcome reports and improvement rates. This causes a tradeoff between the amount of transparency and safety for learning. (Figure 2)

**Figure 2.**
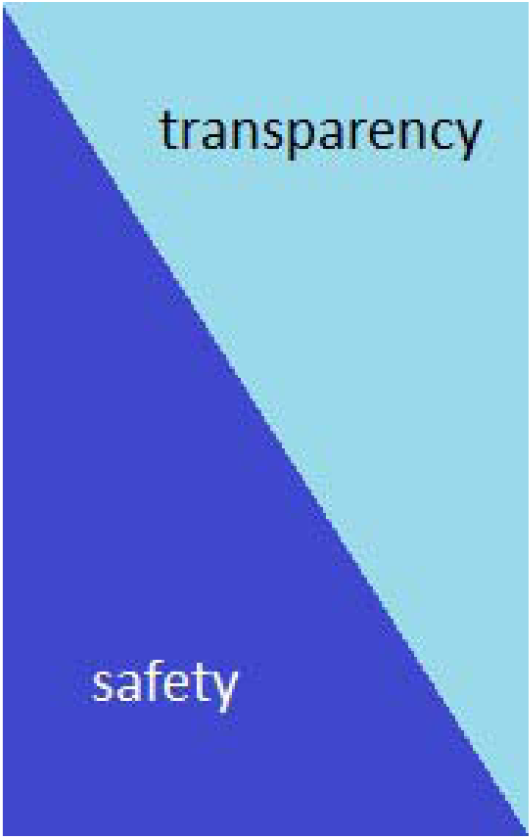
Transparency versus safety for learning

#### Accountability

Sustainable quality improvement depends on health professionals feeling safe enough to learn. At the same time, it is necessary for doctors to realize the importance of offering society insight into their daily practice and their actions to improve it. The era in which accountability was optional is long behind us.^1^ GPs and professional organizations express the desire to take responsibility themselves by developing a culture of holding each other accountable within these groups, but this is not yet the reality, as our participants stated. Patients and representatives of insurance companies express difficulty trusting that this will happen successfully. Even so, as one of our participants pointed out: there are already structures in place to hold poorly performing doctors accountable.

Fortunately, our results show that there is room for conversation on both sides: GPs and professional organizations are aware that there has to be a certain level of accountability. Insurance companies also understand that doctors need to feel safe to make a mistake here and there in order to learn. Meanwhile, patients explain that trusting their doctor does not have much to do with numbers and data, but far more with their personal experience.

Given the above, we urge health care professionals and policy makers to have a conversation on transparency versus a safe learning environment within quality improvement. When designing and introducing new quality improvement measures, awareness of the tension between transparency and a safe learning environment is crucial. Clarity on wanted purposes, learning, and/or assessment should be properly discussed with all stakeholders involved. It should be questioned whether both of these purposes can exist within the same activity. When both are required, the effects on safe learning should be taken into account and, as suggested by Sargeant et al, carefully managed.^35^ Since this tension has been researched widely in medical educational literature, we believe that using a broader scope of theory, especially educational theory, when conducting quality improvement research, may help with gaining insight into the underlying problems and may offer solutions.^36^

### Strengths, limitations and further research

It is important to note that this is an exploratory study, performed in a specific Dutch general practice context. Opportunities and risks identified in this context and by these stakeholders therefore cannot simply be extrapolated to other settings and larger numbers. We chose to focus on the three key stakeholders in our view, excluding other relevant agencies, such as governmental agencies and the Healthcare Inspectorate. Although we believe that the most important opportunities and risks were identified, different insights may be identified when including these agencies. All differences in settings aside, the struggle to develop new types of quality improvement tools and systems is widely shared across contexts and nations. We may learn from each other’s experiences. Moreover, we believe the tension between transparency and the safe learning environment to be relevant to many other quality improvement contexts.

It proved difficult to find GPs who were critical of AF peer discussion, which raises the question of whether our participating GP population was representative and peer discussion of AF is indeed widely embraced, or whether we were unable to escape the academic minded enthusiastic GP when selecting our sample. We did purposively sample GPs in order to find a critical voice as well, adding another interview with a GP. When interviewed, this GP was merely critical of whether all GPs are indeed part of a safe group, and not of the opportunities AF could offer when indeed performed within a safe peer group. Even so, further research is necessary, in order to verify our findings with a larger number of GPs.

## Conclusion

Peer discussion of audit and feedback is a valuable addition to quality improvement in general practice, according to stakeholders. It offers opportunities to engage with AF reports on a deeper level, resulting in learning and leading towards behavior change. It could be part of changing the quality system in general practice towards a system based on learning. Creating a safe learning environment is a key part of this. Since tension exists between learning and improvement in a safe environment on the one hand, and asking for a high degree of transparency on the other, using peer discussion of AF for accountability purposes should be treated with caution.

## Supporting information

final code tree

SRQR-checklist

## Data Availability

All data produced in the present study are available upon reasonable request to the authors.

