## Supplementary material for "Walking the line between assessment, improvement, and learning: an exploratory study on opportunities and risks of peer discussion of audit and feedback": final code tree

### Title page

Word count: 5351

Key words: clinical audit, continuing medical education, formative feedback, general practice, peer group

Code System ‘Walking the Line between assessment, improvement, and learning: an exploratory study on opportunities and risks of peer discussion of audit and feedback’

Code system

Opportunities

Opportunities\reeching deeper levels of reflection

Opportunities\reeching deeper levels of reflection\GPs

Opportunities\reeching deeper levels of reflection\GPs\uncovering blind spots

Opportunities\reeching deeper levels of reflection\GPs\group asking different questions

Opportunities\reeching deeper levels of reflection\GPs\group offering different solution

Opportunities\reeching deeper levels of reflection\GPs\group encouraging to improve

Opportunities\reeching deeper levels of reflection\Insurance companies

Opportunities\reeching deeper levels of reflection\Insurance companies\ group asking different questions

Opportunities\reeching deeper levels of reflection\Insurance companies \group encouraging to improve

Opportunities\reeching deeper levels of reflection\POs

Opportunities\reeching deeper levels of reflection\POs\group discussion as second mirror

Opportunities\reeching deeper levels of reflection\POs\uncovering blind spots

Opportunities\reeching deeper levels of reflection\POs\group asking different questions

Opportunities\reeching deeper levels of reflection\patients

Opportunities\reeching deeper levels of reflection\patients\improving together as a necessity

Opportunities\adding context to numbers

Opportunities\ adding context to numbers \GPs

Opportunities\ adding context to numbers \GPs\complexity of general practice care

Opportunities\ adding context to numbers \GPs\reality is different from numbers

Opportunities\ adding context to numbers \GPs\group adds context

Opportunities\ adding context to numbers \Insurance companies

Opportunities\ adding context to numbers \Insurance companies\indicators are a starting point

Opportunities\ adding context to numbers \Insurance companies\GPs in the lead on meaning of numbers

Opportunities\ adding context to numbers \POs

Opportunities\ adding context to numbers \POs\complexity of general practice care

Opportunities\ adding context to numbers \POs\numbers simplify

Opportunities\ adding context to numbers \POs\group adds context

Opportunities\ adding context to numbers \patients

Opportunities\ adding context to numbers \patients\numbers simplify

Opportunities\taking more ownership

Opportunities\ taking more ownership \GPs

Opportunities\ taking more ownership \GPs\quality improvement is part of the profession

Opportunities\ taking more ownership \GPs\GPs should be in the lead

Opportunities\ taking more ownership \GPs\GPs should be in the lead\ownership prevents constant accountability

Opportunities\ taking more ownership \GPs\GPs should be in the lead\group can mobilize action

Opportunities\ taking more ownership \Insurance companies

Opportunities\ taking more ownership \Insurance companies\quality improvement is part of the profession

Opportunities\ taking more ownership \Insurance companies\we would welcome GPs in the lead

Opportunities\ taking more ownership \POs

Opportunities\ taking more ownership \POs\quality improvement is part of the profession

Opportunities\ taking more ownership \POs\GPs should be in the lead

Opportunities\ taking more ownership \POs\GPs should be in the lead\ownership prevents constant accountability

Opportunities\ taking more ownership \patients

Opportunities\ taking more ownership \patients\trust in GPs

Risks

Risks\How to handle unwilling colleagues?

Opportunities\How to handle unwilling colleagues?\GPs

Opportunities\How to handle unwilling colleagues?\GPs\some people won’t participate

Opportunities\How to handle unwilling colleagues?\GPs\some people won’t participate\they can be excellent doctors

Opportunities\How to handle unwilling colleagues?\Insurance companies

Opportunities\How to handle unwilling colleagues?\Insurance companies\we need some form of control

Opportunities\How to handle unwilling colleagues?\POs

Opportunities\How to handle unwilling colleagues?\POs\some people won’t participate

Opportunities\How to handle unwilling colleagues?\POs\some people won’t participate\their peers know who they are

Opportunities\How to handle unwilling colleagues?\POs\some people won’t participate\group needs to take action

Opportunities\How to handle unwilling colleagues?\POs\some people won’t participate\there are other mechanisms in order to reinforce adequate care

Opportunities\How to handle unwilling colleagues?\patients\we need some form of control

Risks\Lacking a safe group

Risks\Lacking a safe group\GPs

Risks\Lacking a safe group\GPs\most GPs are in a safe group

Risks\Lacking a safe group\GPs\not all groups are safe for learning

Risks\Lacking a safe group\GPs\not all groups are safe for learning\there can be competition

Risks\Lacking a safe group\POs

Risks\Lacking a safe group\POs\safe group should be available

Risks\patients should be involved

Risks\patients should be involved\GPs

Risks\patients should be involved\GPs\most important outcome is quality of life

Risks\patients should be involved\GPs\most important outcome is patient satisfaction

Risks\patients should be involved\GPs\patients could offer insight

Risks\patients should be involved\patients

Risks\patients should be involved\patients\patients are missing in peer quality cycle

Risks\patients should be involved\patients\patients could offer insight

Risks\patients should be involved\POs

Risks\patients should be involved\Insurance companies

Disagreement on transparency

Disagreement on transparency\we need some form of accountability

Disagreement on transparency\we need some form of accountability\GPs should be able to decide on form

Disagreement on transparency\we need some form of accountability\you cannot grade your own exam

Disagreement on transparency\process evaluation

Disagreement on transparency\process evaluation\offering insight in that you participate should be plentiful

Disagreement on transparency\process evaluation\offering insight in how you participate should be plentiful

Disagreement on transparency\process evaluation\besides process eval we need some outcome measurement

Disagreement on transparency\outcome measurements

Disagreement on transparency\outcome measurements\outcome measurements should not be asked

Disagreement on transparency\outcome measurements\some form of outcome measurement should be offered

Disagreement on transparency\outcome measurements\outcome measurement could impair safety of learning

Disagreement on transparency\ambiguity
