## Supplementary material for "Walking the line between assessment, improvement, and learning: an exploratory study on opportunities and risks of peer discussion of audit and feedback": SRQR-checklist

### Title page

Word count: 5351

Key words: clinical audit, continuing medical education, formative feedback, general practice, peer group

**Standards for Reporting Qualitative Research (SRQR)**

O’Brien B.C., Harris, I.B., Beckman, T.J., Reed, D.A., & Cook, D.A. (2014). Standards for reporting qualitative research: a synthesis of recommendations. *Academic Medicine, 89(9)*, 1245-1251.

| **No. Topic** | **Item** | **Page nr** |
| --- | --- | --- |
| **Title and abstract** |  |  |
| S1 Title | Concise description of the nature and topic of the study identifying the study as qualitative or indicating the approach (e.g., ethnography, grounded theory) or data collection methods (e.g., interview, focus group) is recommended | 1 |
| S2 Abstract | Summary of key elements of the study using the abstract format of the intended publication; typically includes objective, methods, results, and conclusions | 2 |
| **Introduction** |  |  |
| S3 Problem formulation | Description and significance of the problem/phenomenon studied; review of relevant theory and empirical work; problem statement | 3,4 |
| S4 Purpose or research question | Purpose of the study and specific objectives or questions | 3,4 |
| **Methods** |  |  |
| S5 Qualitative approach and research paradigm | Qualitative approach (e.g., ethnography, grounded theory, case study, phenomenology, narrative research) and guiding theory if appropriate; identifying the research paradigm (e.g., positivist, constructivist/interpretivist) is also recommended | 4 |
| S6 Researcher characteristics and reflexivity | Researchers’ characteristics that may influence the research, including personal attributes, qualifications/experience, relationship with participants, assumptions, or presuppositions; potential or actual interaction between researchers’ characteristics and the research questions, approach, methods, results, or transferability | 6,7 |
| S7 Context | Setting/site and salient contextual factors; rationale^a^ | 4,5 |
| S8 Sampling strategy | How and why research participants, documents, or events were selected; criteria for deciding when no further sampling was necessary (e.g., sampling saturation); rationale^a^ | 5 |
| S9 Ethical issues pertaining to human subjects | Documentation of approval by an appropriate ethics review board and participant consent, or explanation for lack thereof; other confidentiality and data security issues | 7 / na |
| S10 Data collection methods | Types of data collected; details of data collection procedures including (as appropriate) start and stop dates of data collection and analysis, iterative process, triangulation of sources/methods, and modification of procedures in response to evolving study findings; rationale^a^ | 5,6 |
| S11 Data collection instruments and technologies | Description of instruments (e.g., interview guides, questionnaires) and devices (e.g., audio recorders) used for data collection; if/how the instrument(s) changed over the course of the study | 5,6 |
| S12 Units of study | Number and relevant characteristics of participants, documents, or events included in the study; level of participation (could be reported in results) | 7 |
| S13 Data processing | Methods for processing data prior to and during analysis, including transcription, data entry, data management and security, verification of data integrity, data coding, and anonymization/deidentification of excerpts | 5,6 |
| S14 Data analysis | Process by which inferences, themes, etc., were identified and developed, including researchers involved in data analysis; usually references a specific paradigm or approach; rationale^a^ | 5,6 |
| S15 Techniques to enhance trustworthiness | Techniques to enhance trustworthiness and credibility of data analysis (e.g., member checking, audit trail, triangulation); rationale^a^ | 5,6 |
| **Results/Findings** |  |  |
| S16 Synthesis and interpretation | Main findings (e.g., interpretations, inferences, and themes); might include development of a theory or model, or integration with prior research or theory | 7-14 |
| S17 Links to empirical data | Evidence (e.g., quotes, field notes, text excerpts, photographs) to substantiate analytic findings | 7-14 |
| **Discussion** |  |  |
| S18 Integration with prior work, implications, transferability, and contribution(s) to the field | Short summary of main findings; explanation of how findings and conclusions connect to, support, elaborate on, or challenge conclusions of earlier scholarship; discussion of scope of application/generalizability; identification of unique contribution(s) to scholarship in a discipline or field | 14-18 |
| S19 Limitations | Trustworthiness and limitations of findings | 18 |
| **Other** |  |  |
| S20 Conflicts of interest | Potential sources of influence or perceived influence on study conduct and conclusions; how these were managed | 19 |
| S21 Funding | Sources of funding and other support; role of funders in data collection, interpretation, and reporting | 18 |

^a^The rationale should briefly discuss the justification for choosing that theory, approach, method, or technique rather than other options available, the assumptions and limitations implicit in those choices, and how those choices influence study conclusions and transferability. As appropriate, the rationale for several items might be discussed together.
